# Metagenomic investigation of febrile illness in Pakistan reveals global transmission and co-circulation of Zika and Dengue viruses

**DOI:** 10.1101/2025.03.10.25322803

**Authors:** Najeeha T. Iqbal, Kaitlin Sawatzki, Kumail Ahmed, Jennifer Tisoncik-Go, Elise Smith, Kathleen Voss, John Cornelius, Lu Wang, Alicen B. Spaulding, Leonid Serebryannyy, Daniel C. Douek, Muhammad Asif Syed, Syed Faisal Mahmood, Erum Khan, Wesley C. Van Voorhis, Michael Gale

**Author notes:** Corresponding author: Department of Microbiology and Immunology, 689 23rd Avenue SE, Minneapolis, MN 55455. These first authors contributed equally to this article (Najeeha T. Iqbal, Kaitlin Sawatzki). University of Minnesota (Michael Gale).

## Abstract

We report the first identification of Zika virus in Pakistan following genomic and serological analyses of blood samples from 20 patients with febrile illness. In November 2021, an outbreak of dengue-like illness occurred in the metropolitan city of Karachi. Viral genome capture and sequencing of seven patients revealed six cases of dengue virus serotype 2 and two Zika virus infections, including one dengue and Zika virus co-infection. The next year, following severe flooding, 13 suspected dengue patients were screened by real time qRT-PCR and serology, and 92% (12/13) had evidence of current or recent Zika virus infection. Phylogenetic analyses revealed the Zika viruses originated from Brazil. The most recent observed ancestor dates to 2016, suggesting a prior importation event and ongoing circulation. Our results suggest that Zika virus may be circulating and contributing to disease burden during seasonal Dengue outbreak.

## Introduction

*Orthoflavivirus* is a genus of arthropod-borne, positive strand RNA viruses capable of causing serious disease outbreaks in humans (1). Dengue virus (DENV) and Zika virus (ZIKV) are clinically relevant species with widespread circulation in tropical and subtropical climates via the *Aedes* genus host mosquito species with up to 400 million people at risk of infection each year (2–5). Climate change, travel, trade, and migration contribute to the global spread and circulation of both host and pathogen, increasing pandemic potential and clinical relevance (6–8).

The impact of ZIKV infection on vulnerable populations, such as pregnant women, can have serious consequences. In 2016, the World Health Organization declared a Public Health Emergency of International Concern (PHEIC) in response to the emergence of ZIKV-associated fetal disorders. Congenital Zika syndrome (CZS) is characterized by a wide range of fetal abnormalities at birth, including microcephaly (9). Ongoing physical and developmental delays due to CZS have further been reported in cohort studies and a concurrent surge in the number of travel-associated Zika virus cases prompted concern of global spread (10, 11). The PHEIC demonstrated a critical need for an organized and rapid response against a newly emerged pathogen in a diverse region with a heavy existing burden of arboviral disease (12). Despite decreased transmission rates since 2016, the introduction of ZIKV into new regions and endemic circulation remain a persistent outbreak threat (13).

In 2018, Rajasthan State in India reported the country’s first cases of ZIKV infection (14). In that report, household contacts and unrelated febrile and pregnant individuals near a single index case were tested using a quantitative real time reverse transcription polymerase chain reaction (qRT-PCR) assay. That study found that 7.48% (153/2043) were positive for ZIKV viral RNA, and 40.5% (62/153) were positive among pregnant women. These findings demonstrate that ZIKV can begin circulation in new regions without a specific outbreak event, and that pregnant people may be especially vulnerable to infection.

In November 2021, an unknown viral outbreak in Karachi, Pakistan was reported in local news sources and investigated by the Field Epidemiology Lab Training Program (FELTP) of the Health Department Sindh (15). DENV is endemic to the region and infection outbreaks are common. Here, we report evidence of co-circulation of dengue virus serotype-2 (DENV-2) and Brazil-origin ZIKV associated with this outbreak, as well as additional dengue fever cases in 2022. The observed ZIKV genomes are distinct from those of the Asian-origin virus circulating in neighboring India. This is the first identification of ZIKV in Pakistan. We discuss the implications of previously undetected virus importation, introduction, and circulation.

## Methods

### Study population

Samples were collected from 20 patients with suspected arbovirus disease admitted to Aga Khan Hospital or a private hospital in Karachi, Pakistan between November 2021 and October 2022 (Appendix Figure 1). Enrollment, sample collection and processing protocols were approved by the Ethical Review Committee of Aga Khan University (ERC#4794). The Field Epidemiology Lab Training Program (FELTP) and Aga Khan University Hospital-Pakistan center for United World Antiviral Research Network (UWARN) coordinated sample collection and processing. FELTP is administered by the provincial Health Department in Sindh. UWARN is a multi-institute collaboration under the National Institutes of Health (NIH), National Institute for Allergy and Infectious Disease (NIAID) Centers for Research in Emerging Infectious Diseases (CREID) and is administered at the University of Washington, Seattle, WA, USA.

### Sample collection and processing

Blood samples were collected in EDTA tubes and inverted to mix. Tubes were centrifuged 1000 g for 10 minutes at 4°C and the plasma layer aliquoted. Samples were shipped on dry ice to Pandemic Response Repository through Microbial and Immune Surveillance and Epidemiology (PREMISE) at NIAID for serology. RNA was extracted using the QIAamp Viral RNA Extraction Kit (Qiagen, https://www.qiagen.com) according to the manufacture’s protocol and shipped on dry ice to the University of Washington for sequencing.

### Pan-viral metagenomics sequencing

Samples were stored at −80°C until processing in BSL2+ with BSL3 practices. RNA integrity was evaluated using an Agilent 2100 Bioanalyzer or 4200 TapeStation (Agilent Technologies, https://www.agilent.com). cDNA was generated with Random Primer 6 and the ProtoScript II First Strand cDNA synthesis kit [New England Biolabs (NEB), https://www.neb.com]. The second strand was synthesized using the NEBNext® Ultra™ II Non-Directional RNA Second Strand Synthesis Module (NEB). Library preparation was completed using the Twist Total Nucleic Acids Library Preparation EF Kit 2.0 and Twist UDI Primers (Twist Biosciences, https://www.twistbioscience.com). Bead-purified libraries were checked using the Bioanalyzer High Sensitivity DNA assay (Agilent) and pooled up to 8-plex, with the total pool never exceeding 1500 ng. Indexed pools were hybridized at 70°C for 16 hours with the Twist Comprehensive Viral Research Panel (Twist Biosciences) and amplified using KAPA HiFi HotStart ReadyMix (Roche, https://www.roche.com). Enriched libraries were bead purified, validated with the Bioanalyzer High Sensitivity DNA kit (Agilent) and multiplexed. Samples were sequenced to approximately 10 million total reads using the Illumina NextSeq 500/550 High Output v2 Sequencing Kit (150 cycles) (Illumina, https://illumina.com).

### Viral metagenomics

Raw sequence data were assessed for quality using Illumina’s BaseSpace platform and FastQC (v0.11.8). Sequence adapters and low-quality reads were trimmed with Trimgalore (v0.6.4) and uploaded to Genome Detective (https://www.genomedetective.com)(v2.48)(16). For samples with high confidence DENV or ZIKV calls, trimmed reads were mapped to reference DENV2 (NC_001474.2) and ZIKV (NC_035889.1) using BWA (v0.7.17) and consensus called with Samtools (v1.15.1) and BCFtools (v1.10.2). Viral genome accessions are: (1) ZIKV full genomes, GenBank PQ066186-7; (2) DENV-2 full genomes, GenBank PQ069805-9; and (3) DENV-2, partial genome, GISAID EPI_ISL_19304156. Raw data are available at NCBI SRA SRR32520601-7.

### Phylogenetic analysis

Phylogenetic analysis was performed for assembled Zika (N=2) and dengue 2 viruses (N=6). All human- and mosquito-origin ZIKV and DENV-2 genomes with collection year and continent metadata were collected from BV-BRC (17). Sequences noted as lab or stock cultures, passaged multiple times, duplicates, including >10% Ns or with <90% genome coverage were removed resulting in 3936 DENV2 and 898 ZIKV genomes. Each UTR and gene was aligned using ClustalΩ (v1.2.4), trimmed, and concatenated. IQ-TREE (v1.6.12) (18–20) was used for preliminary maximum likelihood (ML) tree generation. ML trees were iteratively subset to preserve diversity with Treemmer (v0.3) (relative tree length of 90%). West African clade ZIKV was removed as new viruses were inferred to be the Asian clade. The final sequence was trimmed to n=150 DENV2 and n=176 ZIKV genomes (21) (Appendix Figures 2-3).

Appropriateness for molecular clock models was tested on final subset ML trees with TempEst v1.5 using best-fitting root (Appendix Figure 4). For ZIKV, a null set of 1000 randomly scrambled metadata-genomes was processed (GTR+F+R3, ultrafast bootstrap=1001), and the null distribution compared to the observed values (Appendix Figure 5). Clock and tree model selection was performed with BEAST (v1.10.4) using path sampling/stepping stone sampling marginal likelihood estimation with 100 path steps and 1 million chain length (Appendix Tables 1-2). Samples were time-stamped to their collection year with one year uncertainty. Final analyses were independently run 5-6 times on BEAST with a chain length of 200 million generations using a skyline tree prior, relaxed clock with an uncorrelated lognormal distribution, and GTR + 4Γ substitution model with empirical base frequencies. Tracer (v1.7) (22) was used to confirm convergence and ESS values over 200. Trees were combined with LogCombiner v1.10.4, 10% burn-in removed, and maximum clade credibility trees selected with TreeAnnotator v1.10.4. Final trees were visualized in FigTree v1.4. Geographic metadata were integrated with Augur and visualized using Auspice v2.56.0 and ArcGIS Pro v3.2.0.

### Serological assays

Indirect Enzyme-Linked Immunosorbent Assay (ELISA) was performed using the Human Anti-ZIKV IgG ELISA kit (R&D Systems, https://www.rndsystems.com) according to the manufacturer’s recommendations. Plates were read immediately on a SpectraMax Paradigm (Molecular Devices, https://www.moleculardevices.com) at 450 nm with a correction wavelength of 540 nm. Results were normalized by subtracting the background plate OD reading from the assay microplate OD values. A normalized OD > 0.200 was considered a ZIKV-positive sample, 0.100 ≤ OD ≥0.200 as equivocal, and OD < 0.100 as ZIKV-negative.

Electrochemiluminescence assays for IgG against Zika and Dengue 2 NS1 protein were performed using the Meso Scale Discovery platform (MSD, https://www.mesoscale.com). Briefly, 384-well plates were coated overnight at 4°C with 1 µg/mL Zika NS1 or Dengue 2 NS1 (R&D Systems) in 1X phosphate-buffered saline. Plates were washed and blocked for 1 hour with shaking at room temperature. Following blocking, plates were washed and incubated with 1:100 diluted samples and controls for 1 hour with shaking at room temperature. Plates were washed and incubated with 1 µg/mL Sulfo-tag anti-human IgG (MSD) for 1 hour at room temperature with shaking. Plates were washed, 1X MSD read buffer applied, and read on a Meso Sector S600 (MSD). Sample signal from each Zika NS1 readout was divided by Dengue-2 NS1 readout to calculate relative binding enrichment.

### Quantitative real time reverse transcription polymerase chain reaction (qRT-PCR)

Extracted RNA was tested using up to three qRT-PCR assays (Appendix Table 3). 1. Pan-DENV with multiplexed probes for the identification of dengue serotypes 1-4 (Biosearch Technologies, https://www.biosearchtech.com), 2. Pan-ZIKV with a single probe (Biosearch Technologies), and 3. Duplex ZIKV with two probes for distinct amplicons (IDT, https://www.idtdna.com). All primers and probes were diluted to 20 µM and 5 µM, respectively. Real time assays were performed using the Platinum Superscript III Invitrogen One-Step qRT-PCR kit (Thermo Fisher) and 5µL RNA template. qRT-PCR was performed using a CFX96 thermal cycler (Bio-Rad, https://www.bio-rad.com) at 52°C for 15 minutes; 94°C for 2 minutes; 45 cycles at 94°C for 15 seconds, 55°C for 20 seconds (*Acquisition), 60°C for 20 seconds; and a final extension at 68°C for 20 seconds. Each run included a non-template control and positive controls (BEI resources, https://www.beiresources.org).

## Results

### Metagenomic screening of febrile patients during Dengue fever outbreak reveal co-circulation of ZIKV

A local outbreak of suspected dengue fever occurred in Karachi in November 2021 and samples were collected from November to December. Seven patients with symptoms consistent with arbovirus infection including fever, chills, headache and myalgia were selected for metagenomic analysis to identify possible causative pathogens (Appendix Tables 4-5). Viral nucleic acids from patient samples were captured using the Twist Comprehensive Viral Research Panel comprised of over 1 million unique 120bp probes designed to tile the genomes from 15,488 different RNA and DNA viruses. Sequenced reads were analyzed using the Genome Detective platform and several viruses were identified with high confidence, including *pegivirus hominis*, *orthoflavivirus denguei* (Dengue virus 2, DENV2), and *orthoflavivirus zikaense* (Zika virus, ZIKV) (Appendix Table 6). Six patients were found to have DENV2, and two patients had ZIKV, including one individual (patient E) with a co-infection.

Specificity of the Twist pan-viral approach was tested using iterative subsampling and mapping to DENV2 reference genome for positive and negative control patient samples (Appendix Figure 6). The assay and sequencing depth were appropriate for high confidence calls, including differentiation of false positives with low genome coverage. The results were consistent with qRT-PCR assay results, apart from a weakly positive DENV signal (Ct 39.35) that was not identified in patient F metagenomic reads (Table 1).

**Table 1.**
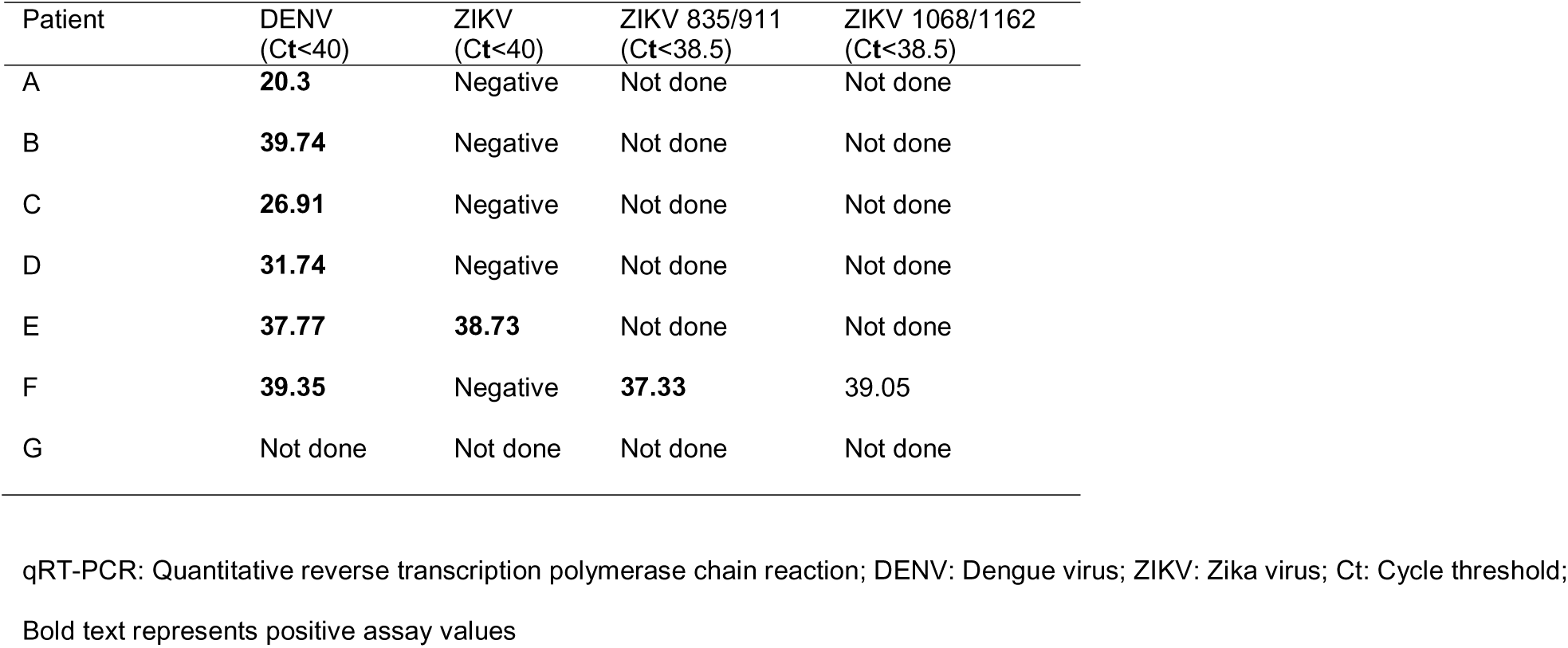
Real time qRT-PCR results for DENV and ZIKV in metagenomics analysis groups.

### Phylogeographic analysis of outbreak-associated DENV and ZIKV

One partial (>90% genome coverage) and seven complete orthoflavivirus genomes were obtained from seven patient samples. No assembled viruses were identical. We performed phylogenetic analyses to estimate the DENV genotype and ZIKV origin. Human- and mosquito-host DENV2 and ZIKV genome sequences with known collection year and source country were obtained from the Bacterial and Viral Bioinformatics Resource Center (BV-BRC) and iteratively pruned from preliminary maximum likelihood trees to retain relevant diversity (Appendix Figures 2-3).

Phylogenetic analysis using a time-aware model in BEAST supports that all observed DENV-2 are DENV-Cosmopolitan genotype of recent East and Southeast Asian origin (Figure 1, Appendix Figure 7). However, the two newly discovered ZIKV strains are more closely related to ZIKV circulating in South America than to contemporaneous ZIKV in neighboring countries (Figure 2, Appendix Figure 8). Phylogeographic inference of virus source using Augur further supports these viruses as an importation event from Brazil to Pakistan (Figure 3).

**Figure 1.**
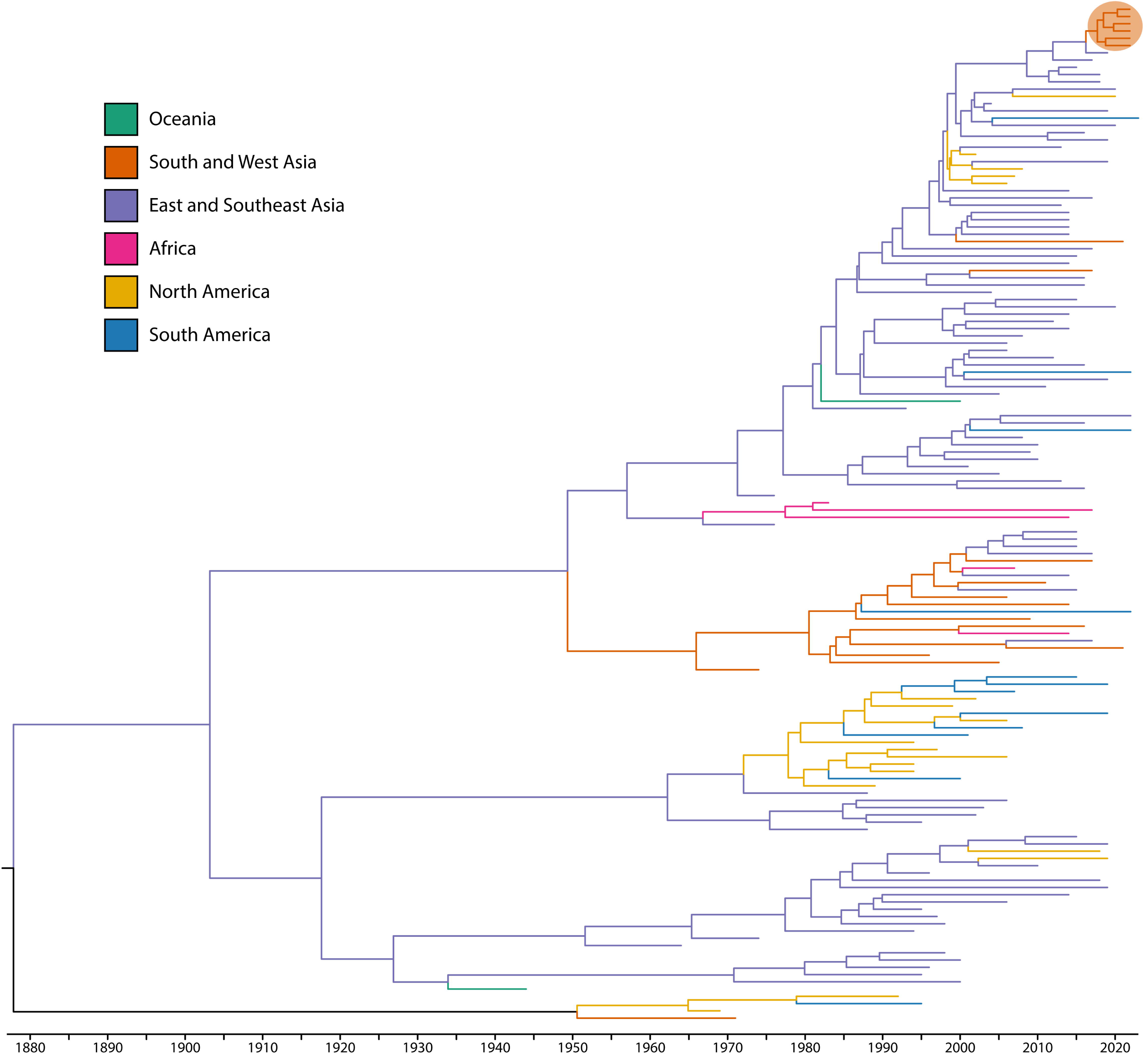
Time-aware phylogeographic tree of patients infected with dengue virus serotype-2 (DENV-2). BEAST time-aware maximum clade credibility tree describing inferred genetic lineage of global DENV, colored by observed and estimated geographic origin. Branch backbones are colored when called with >70% confidence by Augur. Six newly described viruses from Pakistan are circled in orange, with the closest observed ancestors derived from DENV-2 Cosmopolitan genotype in circulating East and Southeast Asia.

**Figure 2.**
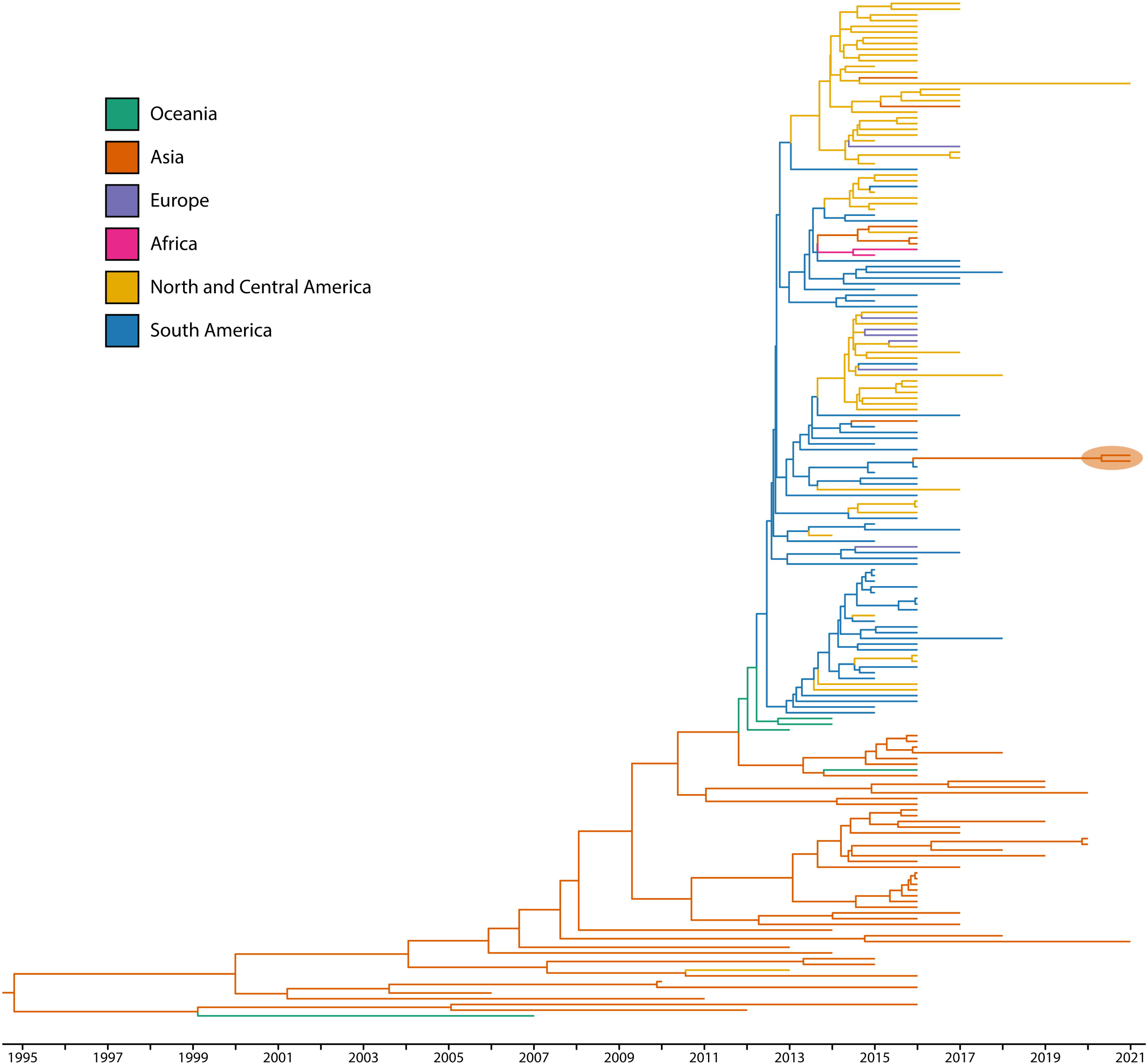
Time-aware phylogeographic tree of patients infected with ZIKV. BEAST time-aware maximum clade credibility tree describing inferred genetic lineage of global Asian-lineage ZIKV, colored by observed and estimated geographic origin. Branch backbones are colored when called with >80% confidence by Augur. Two newly described viruses from Pakistan are circled in orange, with the closest observed ancestors derived from circulating South American ZIKV.

**Figure 3.**
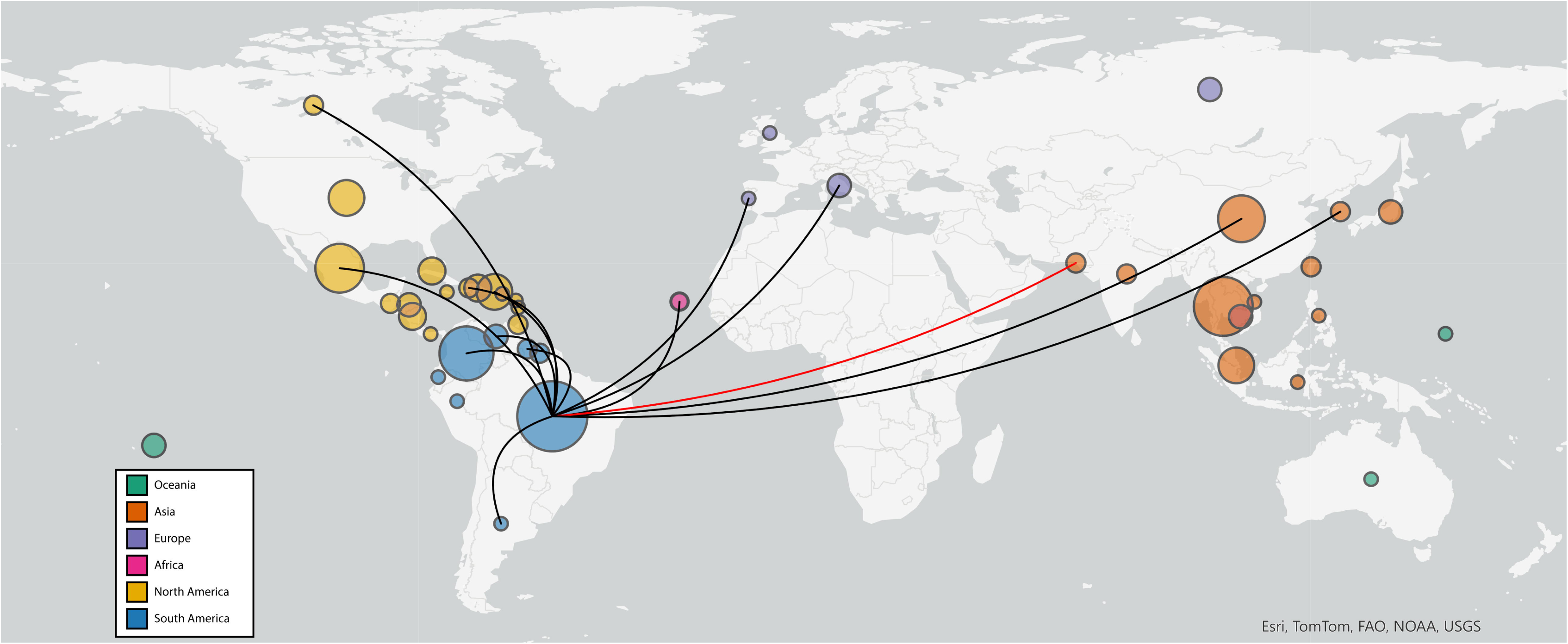
Global map depicting transmission of ZIKV from Brazil to Pakistan. Phylogeographic map illustrating inferred international ZIKV transmission events originating in Brazil. Sequences included in ZIKV phylogenetic tree (Figure 2) were analyzed with Augur and visualized by inferred origin and transmission using Auspice and ArcGIS Pro. Circle size is relative to the number of included viruses from the country and colored by continent. Black lines depict Brazil-origin transmission events in the dataset. Red line highlights the inferred Brazil to Pakistan incursion.

Three ZIKV amino acid changes were unique to the Pakistani viruses and their inferred closest ancestor (KX811222/BRA/2016) as compared to the rest of the Brazilian ZIKV subclade (Figure 4). Two changes are in prM: T74A and S109P. S109P is located in the prM region which maps to the binding interface with Env and both mutations straddle the prM host cleavage site. The third amino acid change is located at the 3’ end of the NS3-coding region, adjacent to NS4A: K587R. There is a final change at the 3’ end of the NS1-coding region encoding M349V that distinguishes the subclade among other Brazilian and South American-circulating viruses (Appendix Figure 9).

**Figure 4.**
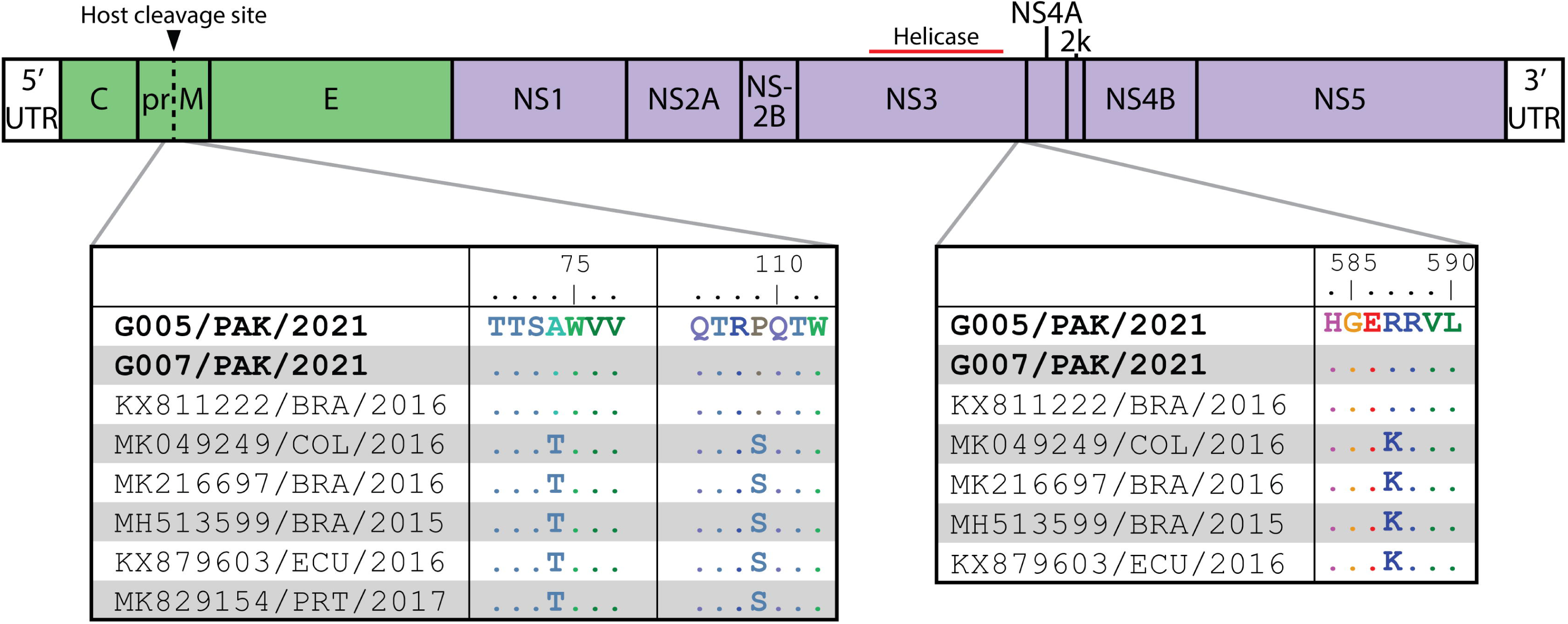
Interclade ZIKV amino acid changes compared to Brazil-origin clade viruses. ZIKV sequences were selected from the same clade and subclade as newly described Pakistan-origin viruses and aligned to G005/PAK/2021. Identical amino acid residues are shown as dots. Two changes were identified in prM, T74A and S109P (left) and one in NS3, K587R (right). G005/PAK/2021 and G007/PAK/2021 correspond to Patients E and F, respectively.

### Viral genome detection and and serological screening of suspected arbovirus cases

Thirteen additional patients were admitted to Aga Khan Hospital from May through October 2022 with febrile symptoms of unknown etiology. We designated these cases as fever of unknown origin (FUO). These patients presented with symptoms characteristic of arbovirus infection, including fever, rash, arthralgia and thrombocytopenia (Appendix Table 7).

Blood samples were collected on days 1 (n=4) and 28 (n=12) following hospital admission and assayed for evidence of active DENV and ZIKV infection by qRT-PCR; ELISA and MSD assays developed by PREMISE (NIAID, NIH) were used to test for serological evidence of recent ZIKV infection via anti-NS1 antibodies (Table 2). All four patients tested positive for DENV by qRT-PCR on day 1. One patient (25%, patient 3) tested positive for ZIKV infection by duplex qRT-PCR assay. Two other patients, not including patient 3, were positive for ZIKV NS1 IgG antibodies. By day 28, 2/12 (17%) were positive for DENV and 0/12 for ZIKV by qRT-PCR assay. However, 10/12 (83%) tested positive for ZIKV IgG by ELISA. Three patients (23%) were antibody positive by both ELISA and MSD (two at day 1, and one at day 28), which strictly controls for potential cross-reactivity between DENV-2 and ZIKV NS1 proteins.

**Table 2.**
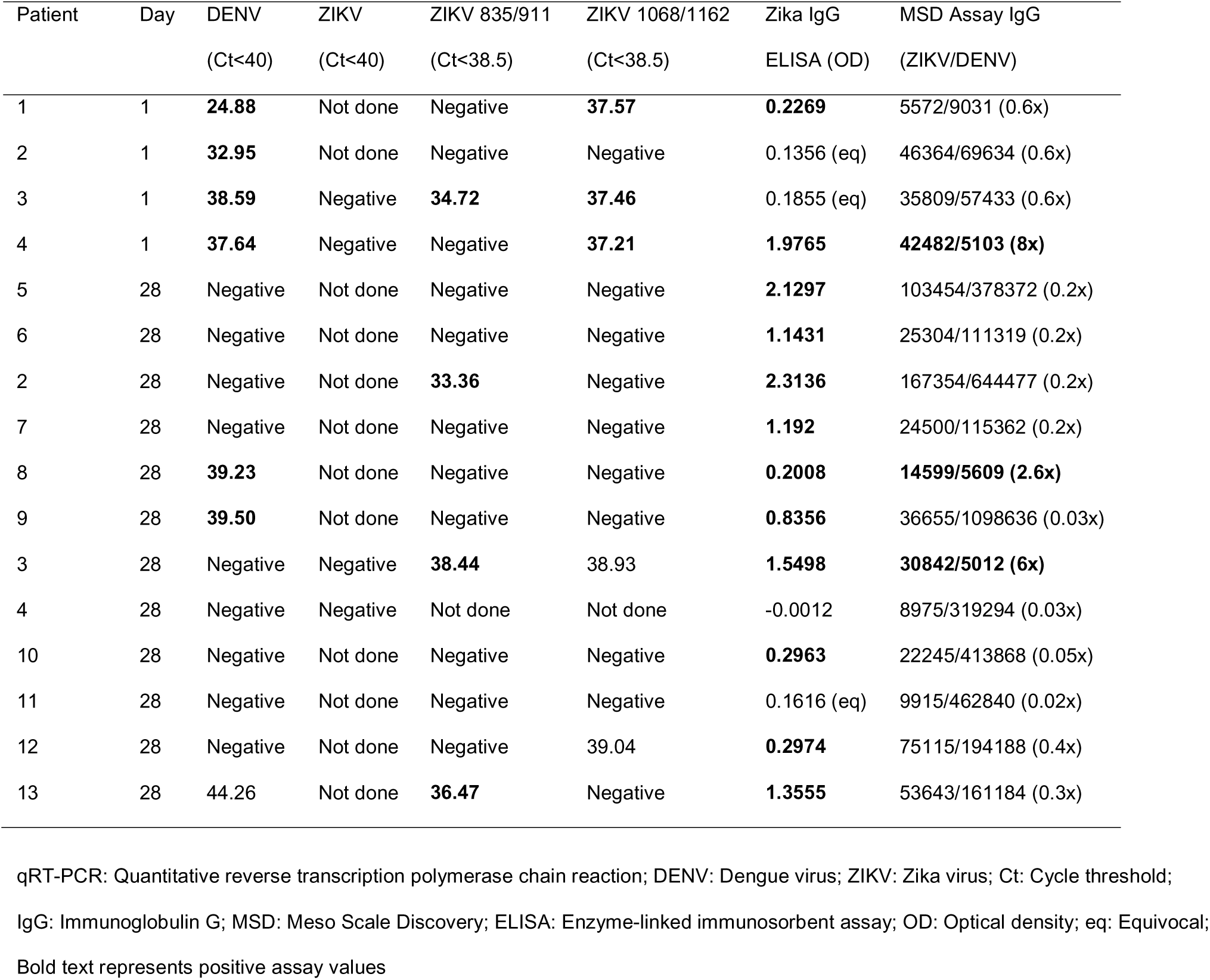
Real time qRT-PCR and serological assay results for DENV and ZIKV infection determination.

### DENV and ZIKV coinfections

Two patients had longitudinal samples consistent with DENV and ZIKV coinfection. The first (patient 3) was symptomatic for seven days upon admission. He presented with respiratory symptoms, high total leukocyte count (22.8×10^9^/L), neutrophil count (88%) and low lymphocyte count (7%). On day 1 of inpatient treatment, he tested positive by qRT-PCR assay for DENV (Ct 38.59) and ZIKV (Cts 34.72, 37.46) coinfection, and was equivocal for ZIKV antibodies (Table 2). By day 28, he developed ZIKV NS1 IgG as measured by ELISA (OD=1.5498) and MSD, which showed a 6-fold change in the ZIKV/DENV-2 NS1 IgG ratio (30842/5012).

The second coinfected individual (patient 4) had been symptomatic for three days upon admission. He presented with suspected dengue fever with a low platelet count (38×10^9^/L), and high lymphocyte count (47.9%). On day 1 of inpatient treatment, he tested positive by both ELISA (OD=1.9765) and MSD (8-fold, 42482/5103) for ZIKV antibodies. The qRT-PCR assay was initially positive for DENV and equivocal for ZIKV (1/2 positive amplicons). By day 28, there was a major peak in the DENV2 NS1 IgG response measured by MSD assay with a corresponding decline in ZIKV IgG (0.03-fold, 8975/319294). These results are suggestive of an initial, symptomatically mild ZIKV infection followed by emergent DENV co-infection. These patients, as well as patient E, from whom we assembled both viral genomes, represent three cases of probable ZIKV-DENV co-infection.

## Discussion

Identification and characterization of etiologic agents associated with infections of unknown etiology in Pakistan is important to understand the consequences of new or re-emerging viruses in the region. Here, we report a pipeline toward virus discovery within specimens of patients presenting with undiagnosed dengue-like illness in the Pakistani province of Sindh. The limitations of traditional antigen and antibody screening can hamper efforts to rapidly diagnose the etiological agent of an outbreak and may miss unusual pathogens. A pan-viral metagenomics assay using the Comprehensive Viral Research Panel offers accessible viral metagenomics on a global scale. We found that this approach can accurately identify viruses with very low read throughput, making it highly scalable by multiplexing.

Using this unbiased discovery approach, we identified ZIKV in Pakistan, and were able to positively identify it in additional samples using a real time qRT-PCR assay. Antibody testing further revealed co-circulation during a dengue fever outbreak, with high seroconversion. ZIKV is an important pathogen for physicians and public health officials to now incorporate in their practice and guidelines.

Arthropod-borne viruses (arboviruses) represent a diverse group of pathogens transmitted primarily by the bite of an infected arthropod, mainly mosquitoes and ticks. In our analysis of ZIKV in Pakistani patients, we uncovered a previously unknown arbovirus importation into the region. Phylogenetic analysis revealed that rather than originating from bordering or near-neighbor countries of Pakistan, the most closely related sequences available originate from Brazil, circa 2016. This distinct clade of Asian-genotype Zika virus emerged in Brazil in 2015 and rapidly spread across the Americas where it is still dominant. Global transmission events are increasingly common in the modern era; Brazil-origin ZIKV from the same time period was exported to many other countries including Canada, Italy, South Korea and Cabo Verde (Figure 3). Most of these events are self-limiting for virus spread, but favorable ecological conditions can establish new areas of virus circulation. The evidence presented in the current study indicates that rather than originating as a singular event, Brazil-origin ZIKV has spread to local *Aedes* mosquitoes and is endemically circulating in Pakistan. ZIKV and DENV overlap in host mosquito species (*Aedes aegypti* and *Aedes albopictus*); therefore, it would be valuable to include ZIKV screening and surveillance in DENV management programs.

Climate change has increased the variation and intensity of monsoon rainfall (23). The samples in this study that were screened by qRT-PCR and serological assays were coincidentally collected during an unprecedented flooding event in Pakistan, including in the Sindh province between July and September 2022. In addition to mortalities, massive displacement and property damage, the 2022 floods caused land degradation which left stagnant water suitable for mosquito habitat. While flooding did not directly affect Karachi, subsequent ecological changes and mosquito-borne disease outbreaks including dengue fever may have contributed to local infections in our study area (24).

Further expansion of surveillance and mosquito control efforts to protect at-risk pregnant populations may be pertinent if CZS cases emerge. As new arboviruses are discovered, outbreaks across diverse, international geographical areas will necessitate the need to interrogate acute and convalescent samples to diagnose causative agents and develop specific diagnostic and therapeutic strategies for use in outbreak responses.

## Data Availability

All data produced are available in the manuscript and online with the following accessions: (1) ZIKV full genomes, GenBank PQ066186-7; (2) DENV-2 full genomes, GenBank PQ069805-9; and (3) DENV-2, partial genome, GISAID EPI_ISL_19304156; (4) Raw data are available at NCBI SRA SRR32520601-7.

## Acknowledgements

These studies were funded in part by National Institute for Allergy and Infectious Diseases, National Institutes of Health grant AI151698 and the Intramural Research Program of National Institute for Allergy and Infectious Diseases, National Institutes of Health. We would like to thank Zack Lindbloom-Brown for information technology infrastructure support.

## Biographical Sketch

Dr. Iqbal is an associate professor and vice chair in the Department of Paediatrics and Child Health, and an associate professor in the Department of Biological and Biomedical Sciences at Aga Khan University Medical College in Karachi. Her research is in infectious disease, primarily tuberculosis and enteric diseases.

