## Appendix for "Metagenomic investigation of febrile illness in Pakistan reveals global transmission and co-circulation of Zika and Dengue viruses"

**Appendix Table 1. DENV model selection in BEAST using PS/SS MLE***

| **Clock** | **Tree** | **PS log MLE** | **SS log MLE** | **Rank** |
| --- | --- | --- | --- | --- |
| Relaxed | Skyline | -112129.573943 | -112135.800803 | 1 |
| Relaxed | Exponential | -112134.208748 | -112137.035396 | 2 |
| Relaxed | Skyride | -112158.659198 | -112166.268221 | 3 |
| Relaxed | Constant | -112169.031368 | -112170.515417 | 4 |
| Random | Skyride | -112241.642808 | -112248.124269 | 5 |
| Random | Skyline | -112250.854750 | -112258.964391 | 6 |
| Random | Exponential | -112444.491463 | -112434.448338 | 7 |
| Random | Constant | -112622.701123 | -112623.790487 | 8 |
| Strict | Skyline | -113240.105197 | -113242.721905 | 9 |
| Strict | Exponential | -113259.065631 | -113257.723317 | 10 |
| Strict | Skyride | -113288.997921 | -113292.263829 | 11 |
| Strict | Constant | -113302.365635 | -113304.192591 | 12 |

*Path sampling (PS)/stepping stone (SS) marginal likelihood estimation (MLE); DENV: Dengue virus; BEAST: Bayesian Evolutionary Analysis Sampling Trees

**Appendix Table 2. ZIKV model selection in BEAST using PS/SS MLE***

| **Clock** | **Tree** | **PS log MLE** | **SS log MLE** | **Rank** | |
| --- | --- | --- | --- | --- | --- |
| Relaxed | Skyline | -41538.701364 | -41544.221456 | | 1 |
| Random | Skyline | -41564.818442 | -41572.615305 | | 2 |
| Relaxed | Constant | -41582.924403 | -41586.979714 | | 3 |
| Relaxed | Exponential | -41591.147698 | -41594.833115 | | 4 |
| Random | Skyride | -41613.561723 | -41615.591782 | | 5 |
| Relaxed | Skyride | -41611.693660 | -41615.931166 | | 6 |
| Random | Constant | -41626.134183 | -41625.191906 | | 7 |
| Strict | Skyline | -41628.226215 | -41635.039381 | | 8 |
| Strict | Exponential | -41648.922299 | -41652.766987 | | 9 |
| Strict | Constant | -41683.036957 | -41685.618307 | | 10 |
| Random | Exponential | -41799.307050 | -41801.812929 | | 11 |
| Strict | Skyride | -41837.651897 | -41842.479273 | | 12 |

*Path sampling (PS)/stepping stone (SS) marginal likelihood estimation (MLE); ZIKV: Zika virus; BEAST: Bayesian Evolutionary Analysis Sampling Trees
**Appendix Table 3. Real time qRT-PCR assay primers and probes**

|  | Sequence (5’-3’) | 5’Fluorophore | 3’Quench |
| --- | --- | --- | --- |
| Pan-Dengue |  |  |  |
| DENV1,2,3-F | CAGATCTCTGATGAACAACCAACG |  |  |
| DENV2-F | CAGATCTCTGATGAATAACCAACG |  |  |
| DENV3-F | CAGATTTCTGATGAACAACCAACG |  |  |
| DENV4-F | GATCTCTGGAAAAATGAAC |  |  |
| DENV1,3-R | TTTGAGAATCTCTTCGCCAAC |  |  |
| DENV2-R1 | AGTTGACACGCGGTTTCTCT |  |  |
| DENV2-R2 | AGTCGACACGCGGTTTCTCT |  |  |
| DENV4-R | AGAATCTCTTCACCAACC |  |  |
| DENV1-Pr | pdCpdUpdCGpdCGpdCGpdUpdUpdUpdCAGpdCApdUApdUA | FAM | BHQ-1 plus |
| DENV2-Pr | pdCpdUpdCpdUpdCGpdCGpdUpdUpdCAGpdCApdUApdU | FAM | BHQ-1 plus |
| DENV3-Pr | pdCpdUpdCpdUpdCApdCGpdUpdUpdCAGpdCApdUApdUpdUG | FAM | BHQ-1 plus |
| DENV4-Pr | pdCpdUpdCApdCGpdCGpdUpdUpdCAGpdCApdUApdU | FAM | BHQ plus |
| Pan-Zika |  |  |  |
| ZIKV-F | CAGCTGGCATCATGAAGAAYC |  |  |
| ZIKV-R1 | CACTTGTCCCATCTTCTTCTCC |  |  |
| ZIKV-R2 | CACCTGTCCCATCTTTTTCTCC |  |  |
| ZIKV-Pr | CYGTTGTGGATGGAATAGTGG | CIV550 | BHQ-1 |
| Zika duplex |  |  |  |
| ZIKV 835 | TTGGTCATGATACTGCTGATTGC |  |  |
| ZIKV-911c | CCTTCCACAAAGTCCCTATTGC |  |  |
| ZIKV-1086 | CCGCTGCCCAACACAAG |  |  |
| ZIKV-1162c | CCACTAACGTTCTTTTGCAGACAT |  |  |
| ZIKV-860-Pr | CGGCATACAGCATCAGGTGCATAGGAG | FAM | BHQ-1 plus |
| ZIKV-1107-Pr | AGCCTACCTTGACAAGCAGTCAGACACTCAA | FAM | BHQ-1 plus |

qRT-PCR: Quantitative reverse transcription polymerase chain reaction; F: Forward; R: Reverse; Pr: Probe

**Appendix Table 4. Summary of patient metadata**

| Patient | Sex | Age | Fever | Chills | Headache | Myalgia/ arthralgia | Nausea | Rash | Hospital diagnosis |
| --- | --- | --- | --- | --- | --- | --- | --- | --- | --- |
| A | F | 46-50 | Y |  | Y | Y | Y | Y | FUO |
| B | M | 16-20 | Y | Y | Y | Y |  |  | FUO |
| C | F | 11-15 | Y | Y | Y | Y | Y |  | FUO |
| D | F | 31-35 | Y |  | Y | Y |  |  | Fever, UTI |
| E | M | 56-60 | Y | Y |  | Y | Y | Y | Dengue fever |
| F | F | 26-30 | Y | Y |  | Y | Y | Y | FUO |
| G | M |  |  |  |  |  |  |  |  |

Y: yes; FUO: fever of unknown origin; UTI: urinary tract infection

**Appendix Table 5. Key study time points for enrolled patients following day 1 symptom onset**

| Patient | Symptom onset | Hospital admission | Sample collection | Hospital discharge | Source |
| --- | --- | --- | --- | --- | --- |
| A | Nov 2021 | Day 7 | Day 9 | Day 11 | FELTP |
| B | Nov 2021 | Day 7 | Day 10 | Day 9 | FELTP |
| C | Nov 2021 | Day 6 | Day 8 | Day 10 | FELTP |
| D | Nov 2021 | Day 4 | Day 9 | Day 9 | AKU |
| E | Nov 2021 | Day 4 | Day 7 | Day 8 | AKU |
| F | Nov 2021 |  | Day 6 |  | AKU |
| G | Nov 2021 |  |  |  | FELTP |

FELTP: Field Epidemiology Laboratory Training Program; AKU: Aga Khan University Hospital

**Appendix Table 6. Top Genome Detective viral metagenomics sequence identification calls per sample**

| Patient | Strain Name | Reads | Depth | Nt ID | Aa ID | Coverage |
| --- | --- | --- | --- | --- | --- | --- |
| A | Dengue virus 2 | 79318 | 3852.39 | 0.94 | 0.97 | 21.51 |
| B | Pegivirus hominis | 1018151 | 19416.41 | 0.90 | 0.97 | 63.92 |
| B | Dengue virus type 2 | 1035353 | 25586.34 | 0.92 | 0.97 | 42.14 |
| C | Dengue virus type 2 | 16600 | 198.32 | 0.92 | 0.97 | 98.79 |
| C | Human endogenous retrovirus K113 | 155 | 11.39 | 0.92 | 0.90 | 16.53 |
| C | Bracoviriform glomeratae | 6 | 4.88 | 0.77 | 0.91 | 31.22 |
| C | Harvey murine sarcoma virus | 2 | 1.37 | 0.84 | 0.92 | 14.24 |
| C | Harvey murine sarcoma virus | 2 | 1.13 | 0.77 | 0.95 | 18.36 |
| D | Dengue virus 2 | 120478 | 1965.41 | 0.92 | 0.97 | 63.25 |
| E | Dengue virus 2 | 51778054 | 565067.79 | 0.92 | 0.97 | 97.42 |
| E | Zika virus | 2956 | 32.71 | 1.00 | 1.00 | 95.32 |
| E | SARS-related coronavirus | 155 | 2.79 | 1.00 | 0.98 | 21.21 |
| E | Sinsheimervirus phiX174 | 28 | 1.38 | 1.00 | 0.99 | 42.80 |
| F | Zika virus | 4573670 | 50188.99 | 1.00 | 1.00 | 95.71 |
| G | Dengue virus 2 | 19125 | 1270.28 | 0.92 | 0.96 | 15.08 |
| G | Betainfluenzavirus influenzae | 1040 | 965.27 | 1.00 | 0.97 | 9.49 |

Nt ID: Nucleotide identity; Aa ID: Amino acid identity

**Appendix Table 7. Sample metadata for patient real time qRT-PCR and serology screening**

| Patient | Age | Gender | Collection month | Days of symptoms | Collection day* |
| --- | --- | --- | --- | --- | --- |
| 1 | 41-45 | Male | May 2022 | 7 | 1 |
| 2 | 51-55 | Male | May 2022 | 3 | 1 |
| 5 | 36-40 | Male | May 2022 | 5 | 28 |
| 6 | 21-25 | Male | May 2022 | 2 | 28 |
| 2 | 51-55 | Male | Jun 2022 | 3 | 28 |
| 3 | 71-75 | Male | Jun 2022 | 7 | 1 |
| 4 | 6-10 | Male | Jun 2022 | 3 | 1 |
| 7 | 26-30 | Female | Jun 2022 | 2 | 28 |
| 8 | 31-35 | Male | Jun 2022 | 3 | 28 |
| 9 | 16-20 | Female | Jun 2022 | 3 | 28 |
| 3 | 71-75 | Male | Jul 2022 | 7 | 28 |
| 4 | 6-10 | Male | Jul 2022 | 3 | 28 |
| 10 | 21-25 | Male | Sep 2022 | 3 | 28 |
| 11 | 51-55 | Female | Sep 2022 | 6 | 28 |
| 12 | 41-45 | Male | Sep 2022 | 2 | 28 |
| 13 | 26-30 | Male | Oct 2022 | 4 | 28 |

qRT-PCR: Quantitative reverse transcription polymerase chain reaction; *Day 1 acute phase, day 28 convalescent phase

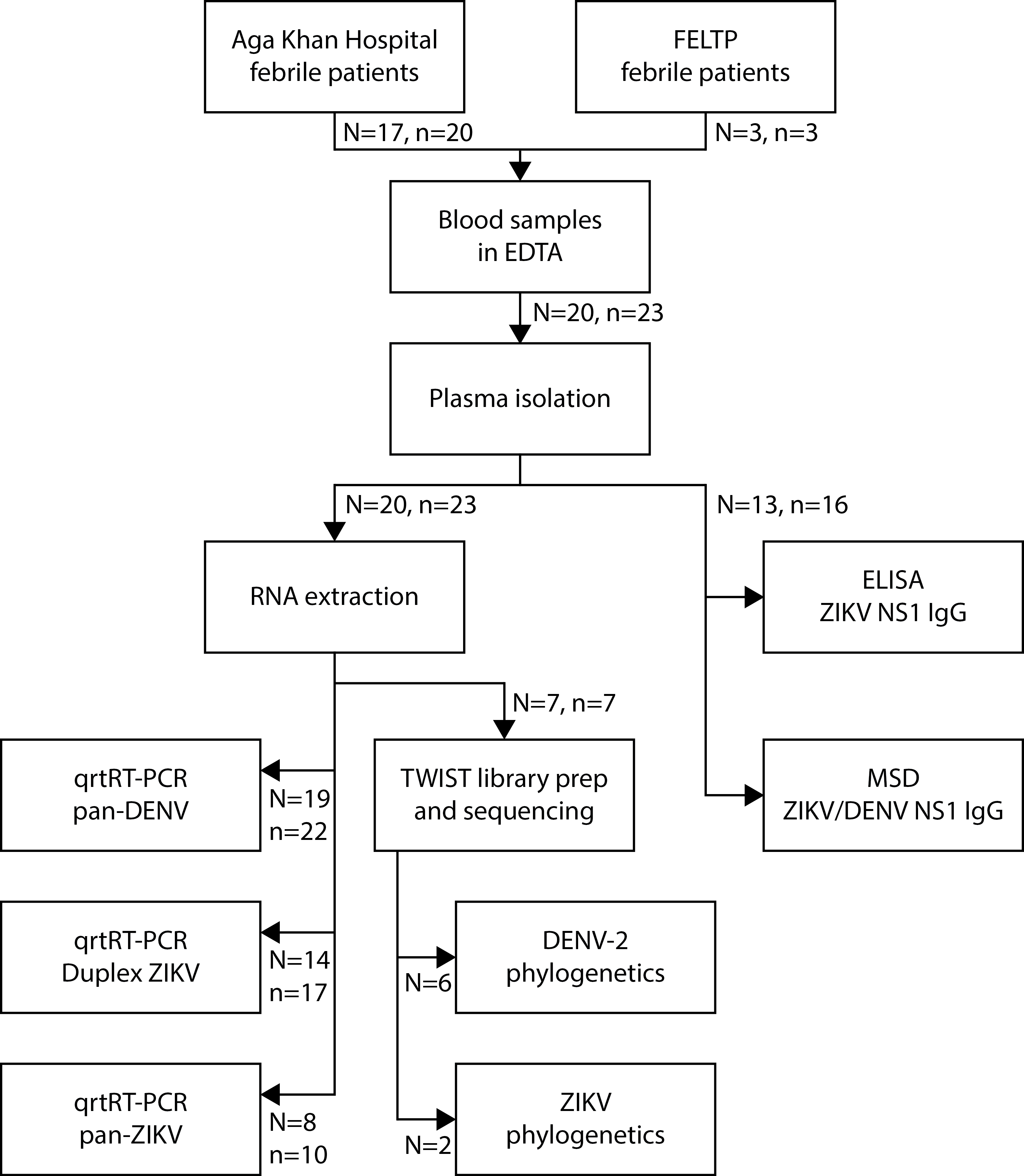

### Appendix Figure 1. Study enrollment and design. Twenty patients presenting with febrile illness at two hospitals in Karachi, Pakistan from November 2021 to October 2022 were enrolled for opportunistic surveillance. Blood was processed to test for (1) evidence of viral RNA using RT-qPCR and sequencing using the Twist pan-viral metagenomis approach, and (2) serologic evidence of recent arbovirus infection by ELISA and MSD in plasma IgG antibodies. N is patient count and n is sample count.

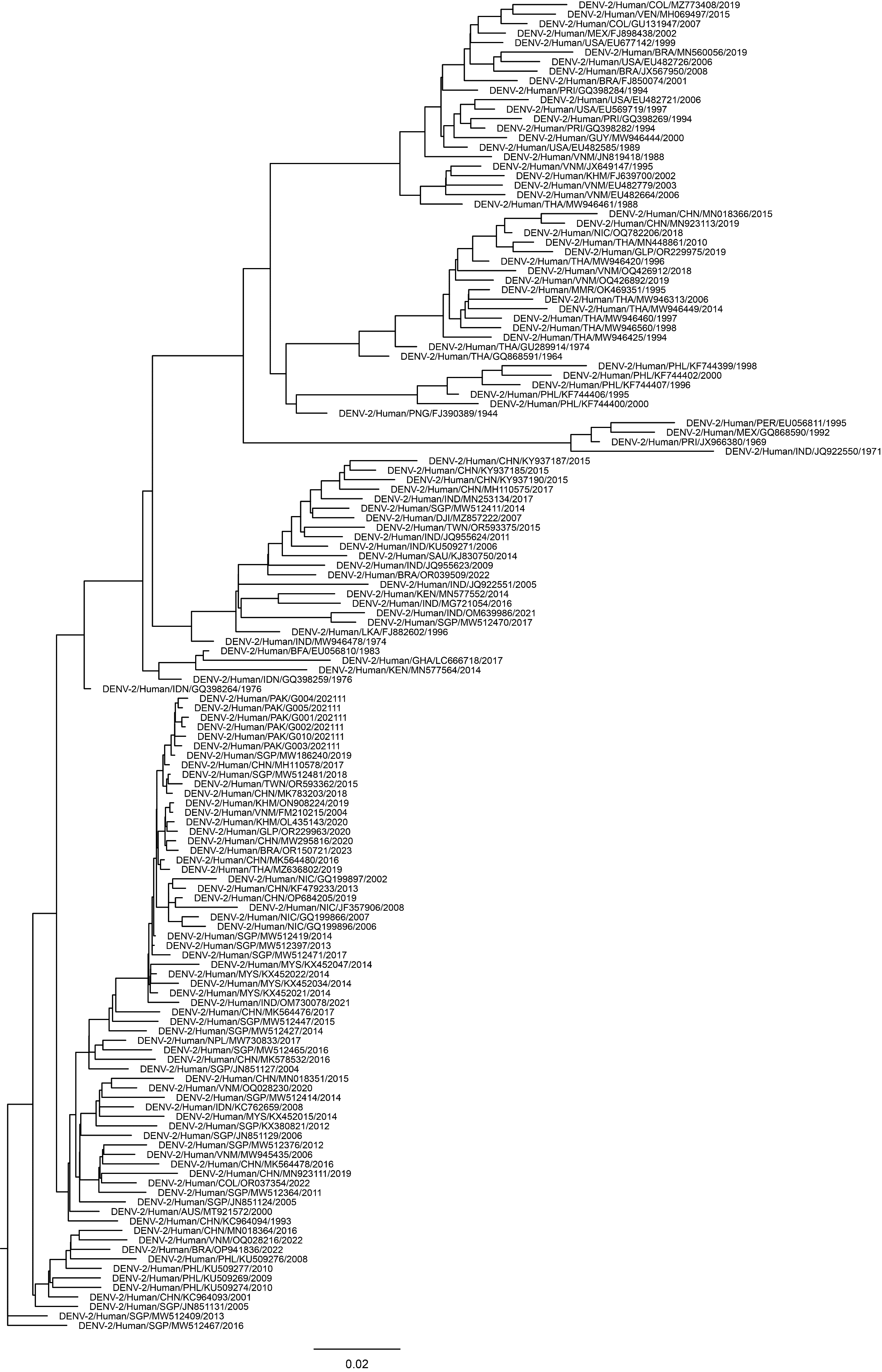

### Appendix Figure 2. DENV-2 maximum likelihood tree. Final DENV-2 sequence set (n=150) phylogenetic tree produced with IQ-TREE using a GTR+F+R3 model with 1001 ultrafast bootstraps.

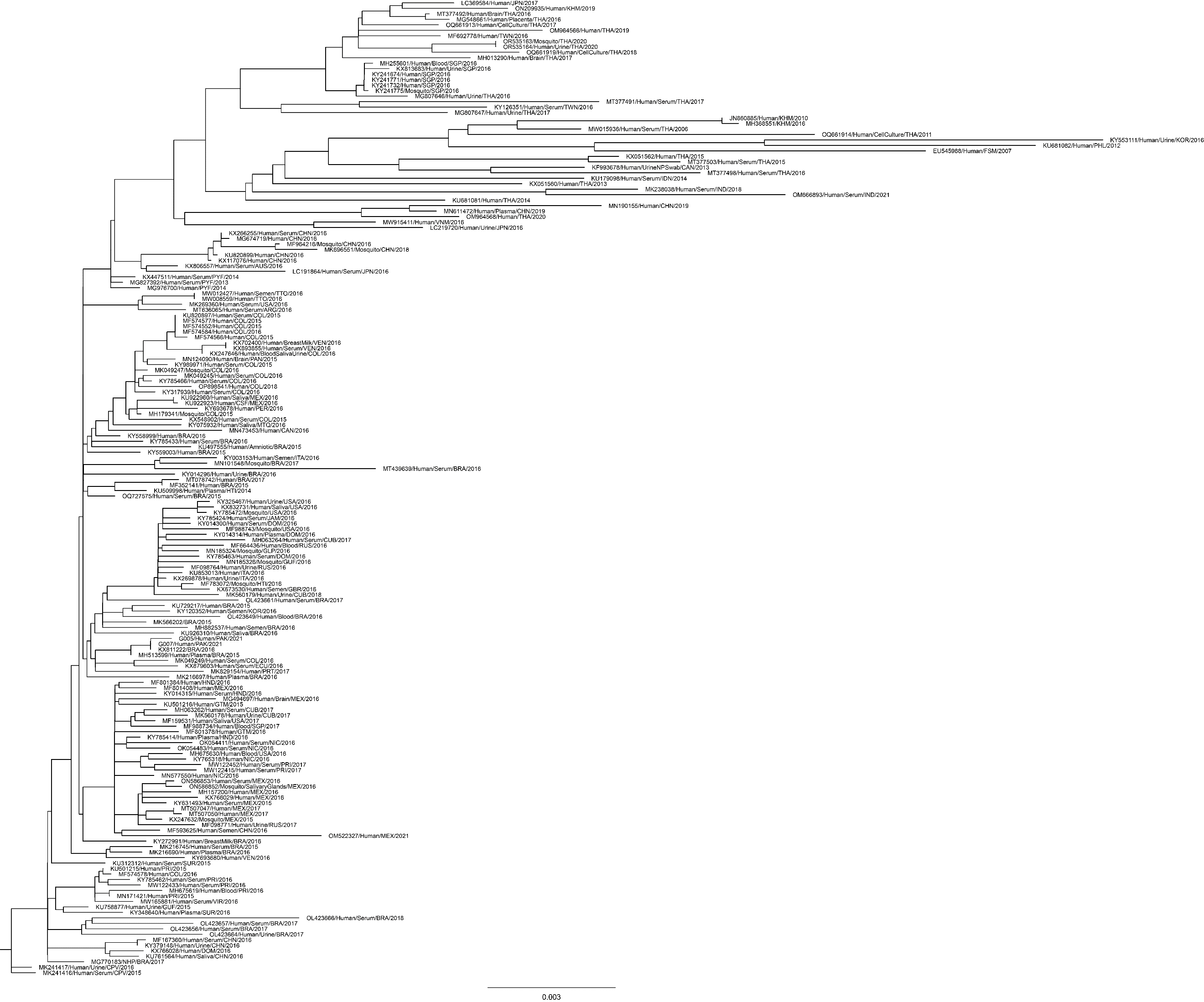

### Appendix Figure 3. ZIKV maximum likelihood tree. Final ZIKV sequence set (n=176) phylogenetic tree produced with IQ-TREE using a GTR+F+R3 model with 1001 ultrafast bootstraps.

A)
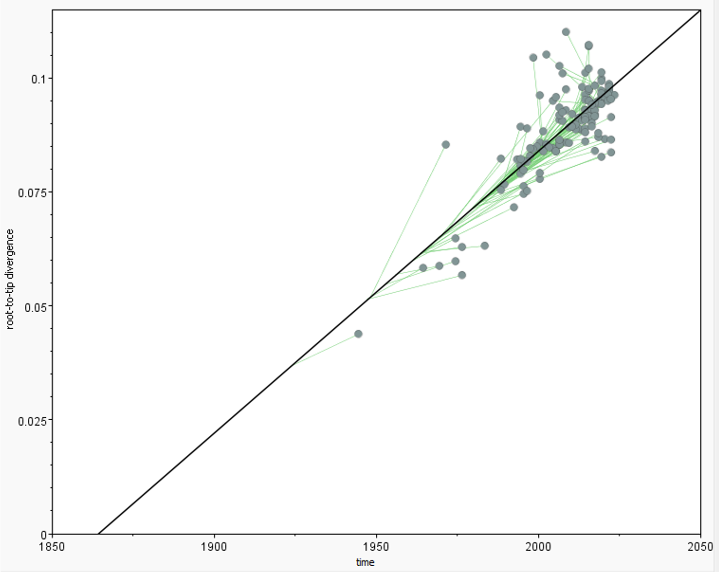

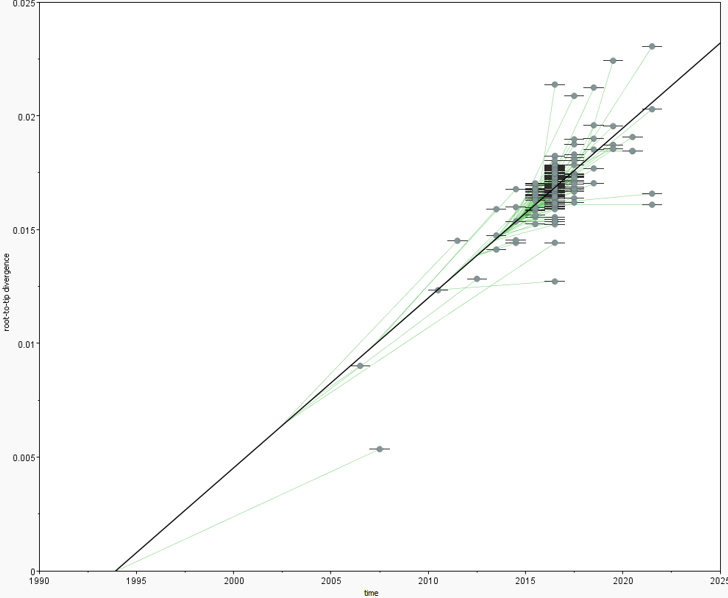
 B)

### Appendix Figure 4. Molecular clock testing. Final maximum likelihood trees for (A) DENV and (B) ZIKV were tested for appropriateness of molecular clock modelling with TempEst using best-fitting root. Correlation coefficients are 0.81 (R2=0.66) for DENV and 0.76 (R2=0.58) for ZIKV.

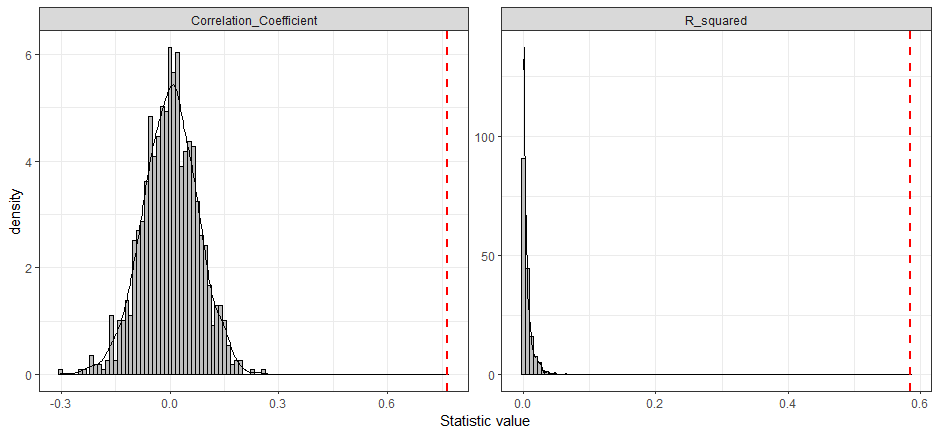

### Appendix Figure 5. Molecular clock testing for ZIKV sequences against a null distribution. A null set of 1000 randomly scrambled genome-metadata was processed through IQ-TREE (m=GTR+F+R3, bb=1001) and TempEst (best-fitting root) and distribution of correlation coefficient and R2 values plotted in grey. Red dashed lines are values of observed data from Appendix Figure 3-4 ML tree.

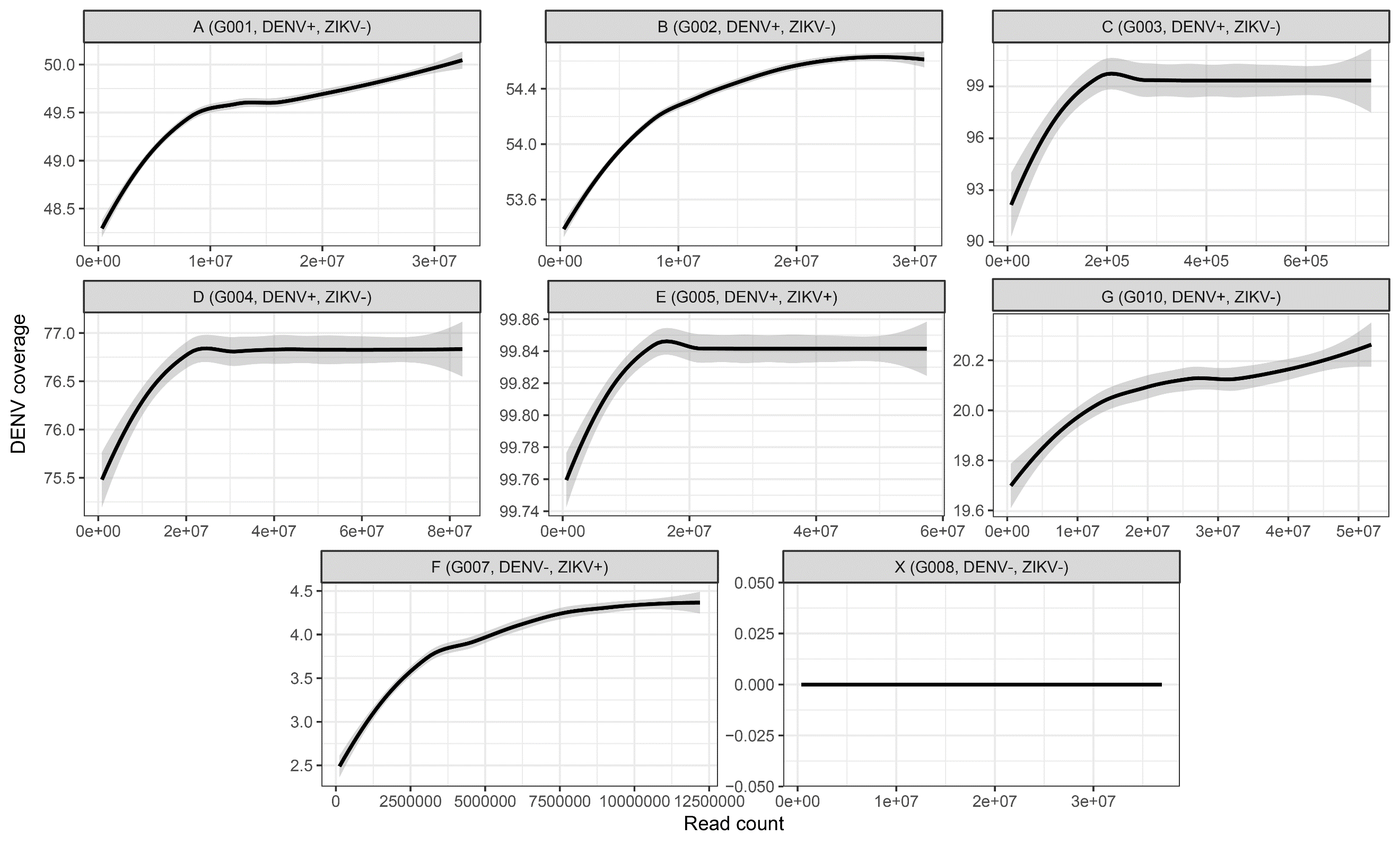

### Appendix Figure 6. DENV genome coverage using TWIST platform. Total reads per sample were mapped to a combined human genome with DENV-1-4 reference. All mapped and unmapped reads were randomly subsampled to represent increasing proportions of total available reads in one percent increments. DENV-2 genome coverage from mapped reads per sample-percent were fit with a LOESS regression and 95% confidence interval shaded in grey. Sample X was prepared in the same manner from a patient without viral fever.

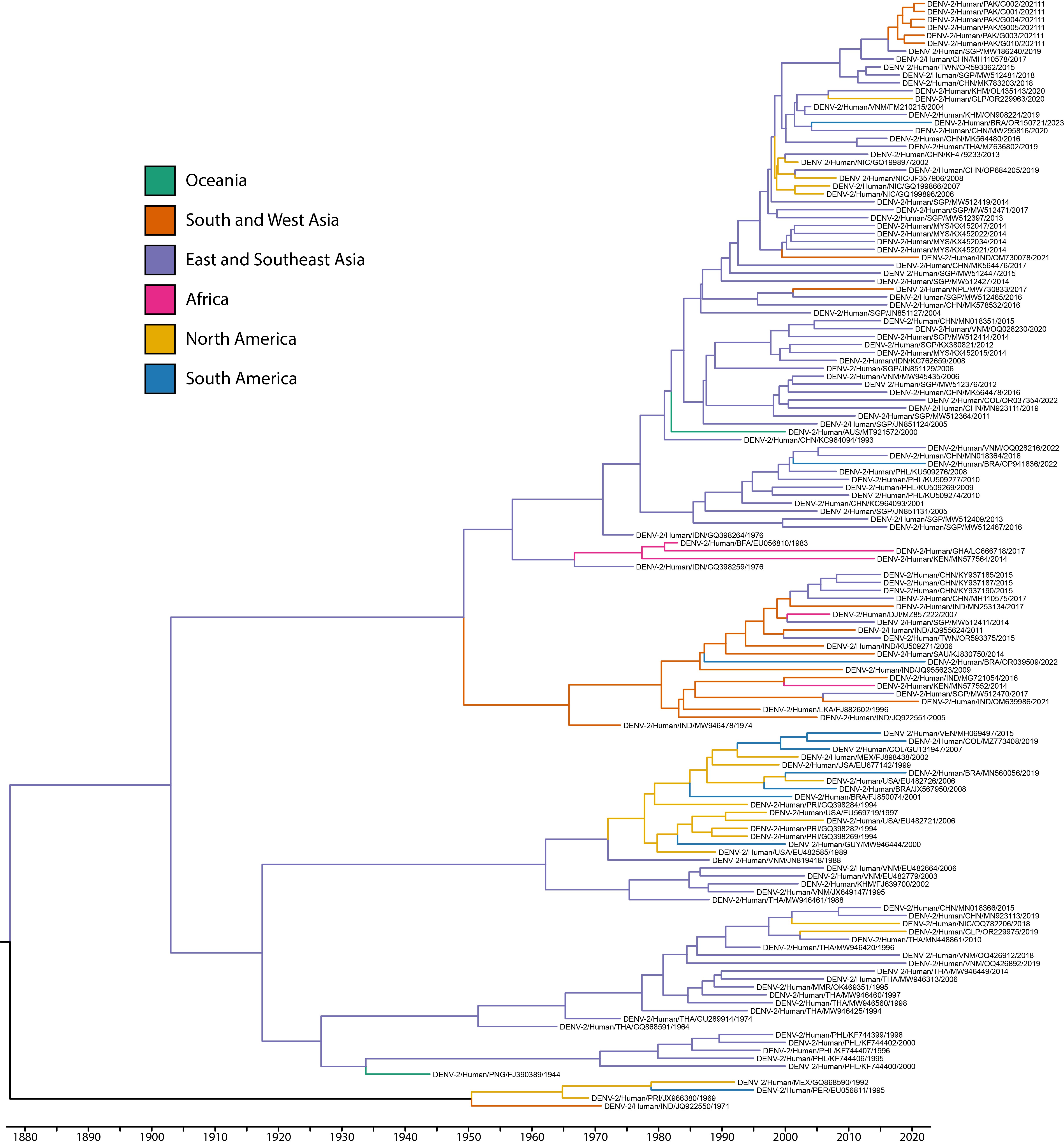

### Appendix Figure 7. Bayesian phylogeographic analysis of patient plasma infected with DENV-2 with sequence metadata. BEAST time-aware maximum clade credibility tree describing inferred genetic lineage of global dengue virus strains, colored by observed and estimated geographic origin. Branch backbones are colored when called with >70% confidence by Augur.

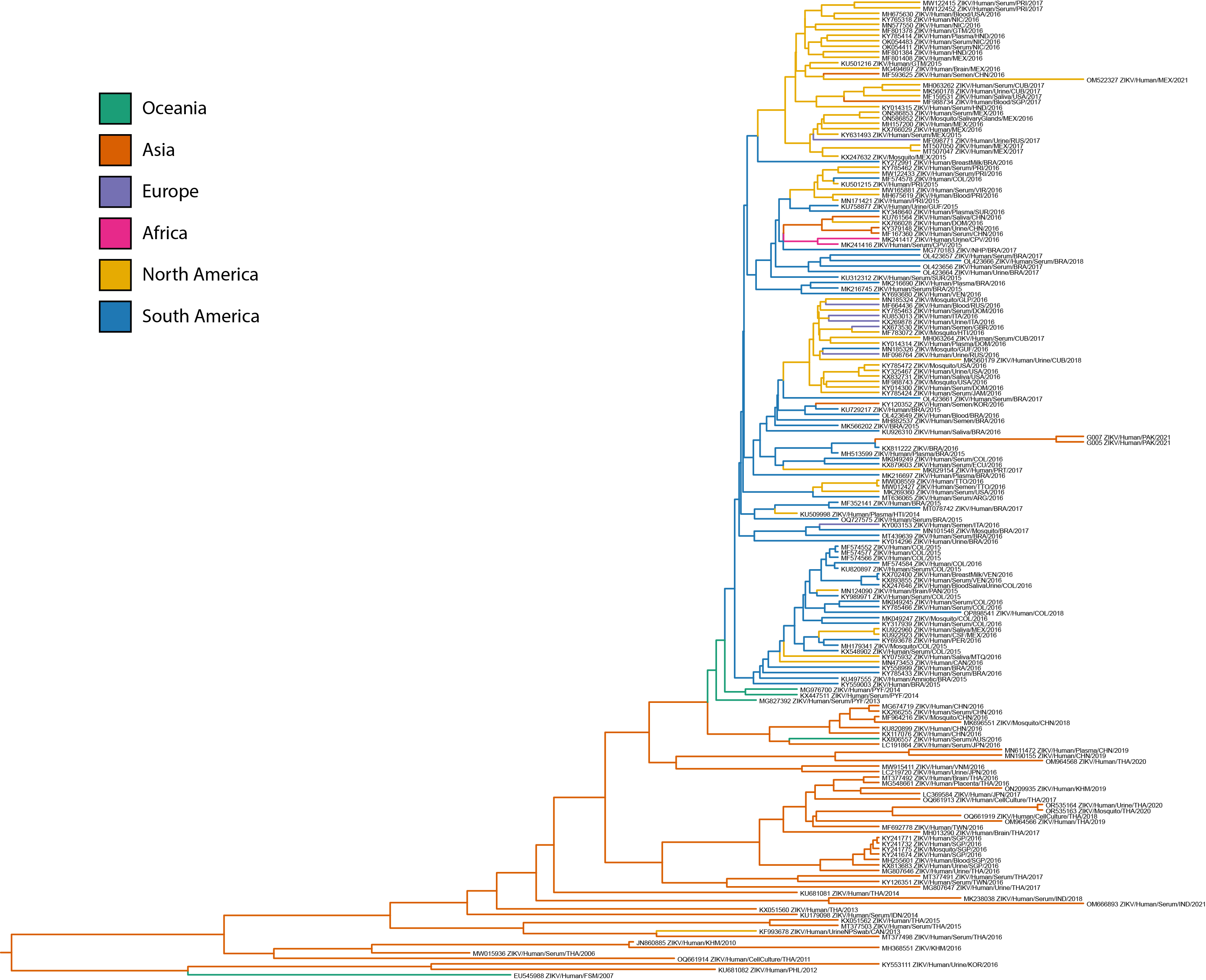

### Appendix Figure 8. Bayesian phylogeographic analysis of patient plasma infected with ZIKV with sequence metadata. BEAST time-aware maximum clade credibility tree describing inferred genetic lineage of global Asian-lineage ZIKV, colored by observed and estimated geographic origin. Branch backbones are colored when called with >70% confidence by Augur.

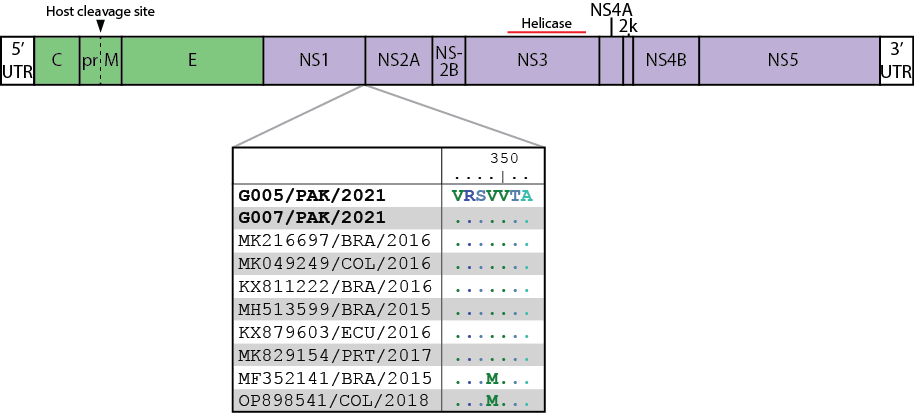

### Appendix Figure 9. Intraclade Zika virus amino acid changes encoded in the NS1 gene. ZIKV sequences were selected from the same clade and subclade as newly described Pakistan-origin viruses and aligned to G005/PAK/2021. Two additional South American-origin viruses were selected outside of the subclade for comparison. Identical amino acid residues are shown as dots. G005/PAK/2021 and G007/PAK/2021 correspond to Patients E and F, respectively. M349V distinguishes the Pakistan-inclusive Brazilian subclade from other South American-circulating viruses.
